# Silent Disease and Loss of Taste and Smell are Common Manifestations of SARS-COV-2 Infection in a Quarantine Facility: Saudi Arabia

**DOI:** 10.1101/2020.05.13.20100222

**Authors:** Alanoud Alshami, Rabab Alattas, Hadeel Anan, Hadi Al Qahtani, Abdulbari Alahilm, Mobarak Al Mulhim, Ahmed Alfaraj

**Affiliations:** Department of Epidemiology and Biostatistics, King Khalid Medical City Research Center, King Fahad Specialist Hospital-Dammam; Division of Immunology, Department of Pathology and Laboratory Medicine, King Fahad Specialist Hospital-Dammam; Divison of Infectious disease, Department of Pediatrics, King Fahad Specialist Hospital-Dammam; King Faisal University, Al-Ahsa, Kingdom of Saudi Arabia; Department of Emergency Medicine, King Fahd Specialist Hospital-Dammam; Department of Pediatrics, King Fahad Specialist hospital-Dammam

## Abstract

**BACKGROUND:** The Coronavirus disease 2019 (COVID-19) outbreak in Saudi Arabia was first identified in a traveler from Al Qatif city, on March 2^nd^, 2020. The disease has quickly spread and reached multiple cities within a few weeks. In an attempt to limit the spread of COVID-19 in Saudi Arabia, the government has implemented strict regulations. Starting March 15^th^, all travelers coming back to the kingdom were tested for COVID-19 and were quarantined in a government-designated facility. The same rule was applied to all positive cases identified by contact tracing. In this study, we aimed to assess the prevalence of asymptomatic carriers, epidemiological characteristics, clinical presentations, and viral clearance of SARS-COV-2 positive quarantined individuals in a quarantine facility in the eastern province.

**METHODS:** We conducted a cross-sectional study on 128 laboratory-confirmed COVID-19 subjects who were quarantined in a government-designated facility. The study period was from March 16^th^ – till April 18^th^, 2020. We collected data on demographics and on clinical symptoms. Also, samples for PCR tests were collected upon admission and were repeated every 72 hours if they were still positive. All negative samples were repeated within 24 hours for confirmation.

**RESULTS:** Sixty-nine of the 128 residents (54%) were completely asymptomatic until the end of the study. The remaining 59 residents (46%) had only mild symptoms. The most common symptom was a sudden loss of smell and taste, accounting for 47.5%. The median time to virus clearance was significantly different between the two groups. Symptomatic residents cleared the virus at a median of 17 days (95% CI,12.4-21.6) from the first positive PCR vs. 11days (95% CI, 8.7-13.3) in the asymptomatic group (P=0.011). False-negative test results occurred in 18.8% of the total residents and false positive results in 3%.

**CONCLUSION:** The prevalence of asymptomatic carriers is high in our study. Testing, and isolating travelers and contacts of laboratory-confirmed cases, regardless of symptoms, were very effective measures for early disease identification and containment. Loss of taste and smell was the most common presentation in our mild symptomatic residents, and it might be predictive of mild disease. The persistent positive PCR beyond 14 days observed in the mild symptomatic residents despite being symptoms free, warrant further studies to determine its implications on disease spread and control.

## Introduction

A novel coronavirus, Severe Acute Respiratory Syndrome coronavirus 2 (SARS-COV-2) was first discovered on December 31^st^, 2019, in Wuhan City, China, after a cluster of atypical pneumonia was observed. This infection was labeled as Corona Virus Disease 2019 (COVID 19)^1^. This has led to an outbreak of infection in China that spread globally over a few months and was declared as a pandemic by the WHO on March 11^th^,2020. Currently, most reported cases are from outside China. As of May 4^th^, 2020, there are 3, 523, 121 confirmed cases worldwide, and 28,656 of these cases are in Saudi Arabia, with 191 reported deaths.

Although we have gradually gained some insight into the clinical presentation spectrum of COVID-19 from other countries' experiences and published data, we still do not have enough information about the real spectrum of the disease and the prevalence and clinical characteristics of asymptomatic carriers. Goa et al. have conducted the first and largest COVID-19 epidemiological study in China. Their study included 72, 314 subjects and they described a mild disease course in about 80% of their cohort, severe disease in 14%, critical in 5%, and asymptomatic in only 1.2%.^2^

The possibility of developing asymptomatic infection has further been reported in larger percentages in multiple small reports ^3,4^, however large data is still lacking. Hu et al. have examined 24 asymptomatic infected individuals with a history of close contact with SARS-COV-2 confirmed cases and found that only 20% of them developed symptoms.

The importance of detecting asymptomatic carriers relies on their ability to spread the disease. This assumption has been illustrated by different reports^4,6^. However, it has not been validated yet in large scale studies. The fact that there is a group of asymptomatic carriers who are roaming around unrecognized and might be able to transmit the disease imposes a serious public health threat if not early identified and contained. Identifying the asymptomatic ones early on will help us better understand the dynamics of the disease spread and guide us to find better measures of disease containment and mitigations.

In this study, we aimed to study the prevalence of true asymptomatic carriers and their clinical characteristics and viral clearance in quarantined individuals with COVID-19.

## Methods

### Study Design and Population

This study was approved by the Institutional Review Board at King Fahad Specialist Hospital-Dammam. This is a retrospective cross-sectional study that was conducted in a quarantine facility in Al-Khobar City in the Eastern province of Saudi Arabia. The facility was designated only for SARS-COV-2 positive cases confirmed by Real-Time Polymerase chain reaction (RT-PCR) using combined nasopharyngeal and oropharyngeal samples. Residents were either traveler who tested positive and transferred from another quarantine, confirmed cases with mild disease transferred from hospitals or mild confirmed cases admitted directly from the community after being traced by the regional public health authority. All residents who were admitted from March 16^th^ until April 6^th^, whether adults or children were included. Admission to the quarantine was restricted to low-risk stable patients with mild symptoms only. Elderly patients >65 years of age with more than one comorbidity were not allowed in this quarantine. All residents were followed up daily by nurses with symptoms screening checklist. Following up, residents on daily bases have allowed us to differentiate between 3 categories of patients: true asymptomatic carriers (no symptoms at all), pre-symptomatic (developed symptoms in the quarantine), and symptomatic patients who had symptoms during or before admission.

### Variables

All data was collected upon presentation to the quarantine. Data was collected prospectively by nurses using the standardized case report forms, generated by modified WHO/ International Severe Acute Respiratory and Emerging Infection Consortium case record form for severe acute respiratory infections. All data collected was verified by the research team through a short phone survey to ascertain the accuracy of the data, especially the symptoms presented within the last 14 days before admission to the quarantine. The collected data included a symptoms-based screening checklists, such as new onset of cough, sore throat, runny nose, fever, and myalgia. Atypical symptoms like headache, nausea and vomiting, diarrhea, and loss of taste and smell. History of comorbidities was taken by self-reported medical history, and it included: Hypertension, Diabetes Mellitus, chronic lung disease, renal disease, and cardiovascular disease. History of pregnancy and current smoking status were also recorded.

PCR samples were taken for each resident upon admission and were repeated every 72 hours as long they were positive. These samples were sent to the regional laboratory for processing, and the results were uploaded on HESN database (Saudi Public health electronic database). Patients were deemed infection-free if they had two negative PCRs 24 hours apart (as per Saudi CDC guideline). We also calculated the percentage of false negative, and false positive tests. A false-negative test was defined as a negative test that was preceded and followed by a positive test within 24-72 hours’ time frame. A false-positive result was defined as a positive test that was preceded and followed by a negative test within 24-72 hours’ time frame.

### Analysis

We mainly used descriptive statistics. Quantitative/continuous variables were presented as mean ± standard deviation (SD) or median with inter-quartile range (IQR). Frequencies, proportions were used to describe qualitative data. Bivariate analysis was performed using an independent sample t-test or Mann-Whitney U test, which is appropriate. Survival analysis was used to determine the time from the first positive PCR until the first true negative one.

### RT-PCR test and sampling

Detection of SARS-CO-V-2 in respiratory samples was performed by RT-PCR using the Altona Kit (RealStar SARS-CoV-2 RT-PCR Kit) (USA). The kits include detection of E gene sequence specific for all B-Beta-corona virus and SARS-COV-2 specific sequence of S gene.

Samples for PCR were collected simultaneously from the nasopharynx and oropharynx following the WHO instructions for sample taking^7^. Each set of the test was reported as screening and confirmatory. RNA was extracted and reverse-transcribed to cDNA. Then real-time PCR was performed as instructed by the manufacture using specific primers and probes targeting the SARS-COV-2 genomes. Internal control is included in the assay to rule exclude reaction inhibition.

### Results

### Epidemiology and Clinical Characteristics

Among the 128 patients included in the study, 69 (53.9%) were female **Table. 1**. The mean age was 39.6 years with a range of (8-76 years). Among the 128 residents, four children <16 years were included. Three out of the four children were asymptomatic, and one child presented with mild fever and cough. Sixty-five residents (50.8%) had a history of travel outside the kingdom within the last 14 days before admission to the quarantine. Iran and Iraq being the most visited countries accounting for (68.2%) followed by UK (10.61%) and the rest of countries accounting for the remaining 21.2 %. Sixty-two residents (49.2%) had a history of direct exposure to a lab-confirmed case, and one resident was screened based on symptoms and tested positive without a clear history of travel or contact with a suspected or lab-confirmed case.

**Table 1.** Demographic Characteristics and Reporting Symptoms of PCR positive quarantined Residents

| Variables | Result<br>(n=128) |
| --- | --- |
| Age ($\bar{x} \pm SD$) —yr. | 39.6 ± 15.5 |
| Gender ( F) — no (%) | 69/128 ( 53.9%) |
| Exposure |  |
| Travel to an effected country | 66 (51.2%) |
| Contact with laboratory confirmed case | 62 (48.8%) |
| Symptoms (n=59) |  |
| Fever >100.4F (38C) | 19 (32.2%) |
| Subjective fever (felt feverish) | 18 (30.5%) |
| Chills | 13 (22.0%) |
| Muscle aches (myalgia) | 23 (39.0%) |
| Runny nose (rhinorrhea) | 17 (28.8%) |
| Sore throat | 20 (33.9%) |
| Cough (new onset or worsening of chronic cough) | 24 (40.7%) |
| Shortness of breath (dyspnea) | 13 (22.0%) |
| Nausea or vomiting | 3 (5.1%) |
| Headache | 22 (37.3%) |
| Abdominal pain | 2 (3.4%) |
| Diarrhea (3 loose/looser than normal stools/24hr period) | 9 (15.3%) |
| loss of tasting and smelling | 28 (47.5%) |
| Other | 7 (11.9%) |
| Chronic disease (n=128) |  |
| Chronic Lung Disease (asthma/emphysema/COPD) | 3 (2.3%) |
| Diabetes Mellitus | 17 (13.2%) |
| Hypertension | 15 (11.6%) |
| Cardiovascular disease | 4 (3.1%) |
| Chronic Liver disease | 1 (.8%) |
| Neurologic/neurodevelopmental/intellectual disability | 3 (2.3%) |
| Current smoker | 20 (15.5%) |
| Former smoker | 3 (2.3%) |
| If female, currently pregnant | 1 (.8%) |
| Other chronic diseases* | 14 (10.9%) |

\* Other chronic disease include: Sickel Cell Anemia, G6PD, and hypothyroidism

69 patients (54.3%) did not exhibit any symptoms (asymptomatic carriers/ silent disease), while the other 59 patients (45.7%) had only mild symptoms.

The most common reported symptom among our residents was a complete and sudden loss of smell and taste, 28 (47.5%). These symptoms appeared at a median of 6 days (IQR, 4-9 days) after the onset of fever or upper respiratory tract symptoms. However, it was the only clinical symptom in 18.65% of patients. Residents also reported a history of cough in (40.7%), myalgia in (39%), and headache in (37.3%). The median time for symptoms resolution was five days (IQR 3-11days).

Out of the 59 symptomatic patients, 11(18.64%) were pre-symptomatic, who subsequently developed symptoms at a mean of 7.78 days (+/- 5.7 SD) after the first positive PCR while in quarantine. The median incubation period in symptomatic residents was 7 days (IQR,3.75-12 days).

### RT-PCR data result

A total of 664 PCR tests have been done for the 128 residents. The median number of tests per person was 5 (IQR, 4-6 tests). Twenty-four residents had at least one false negative test (18.8%) **Table .2**. Symptomatic and asymptomatic residents were significantly different in the timing of clearing the virus. The median time to develop a true negative PCR (2 negative PCRs 24 hours apart) in the symptomatic group was 17 days (95% CI,12.38-21.6) vs. 11 days (95%. CI, 8.66-13.34) in the asymptomatic group (p=0.01)**Figure.1**.

**Table 2.** Virus clearance and swab results with demographic data of the patients.

| Variables | Symptomatic |  | P-value |
| --- | --- | --- | --- |
|  | Yes<br>59 (46.1%) | No<br>69 (53.9%) |  |
| Gender |  |  |  |
| Male | 27 (45.8%) | 32 (45.7%) | 0.999 |
| Female | 32 (54.2%) | 38 (54.3%) |  |
| Age ($\bar{x} \pm SD$) –years | 37.20±13.25 | 41.05±17.59 | 0.159 |
| Cause of Quarantine |  |  |  |
| History of Travel | 22 (37.3%) | 44 (62.9%) | 0.005 |
| History of Contact with<br>lab Confirmed case | 37 (62.7%) | 25 (36.23%) |  |
| Duration in quarantine<br>($\bar{x} \pm SD$) | 16.8± 9.2 | 12.7± 6.2 | .01 |
|  |  |  | . |
| Median time to virus<br>clearance (95% CI) | 17 (12.4 -21.6) | 11(8.7-13.3) | .001 |
| False negative results | 11/59 (18.64%) | 14/69 (20.3%) | .497 |
| False positive results | 5/59 (8.5%) | 4/69(5.8%) | .566 |

Results are expressed as mean ± standard deviation, and number (%).

**Figure.1.**
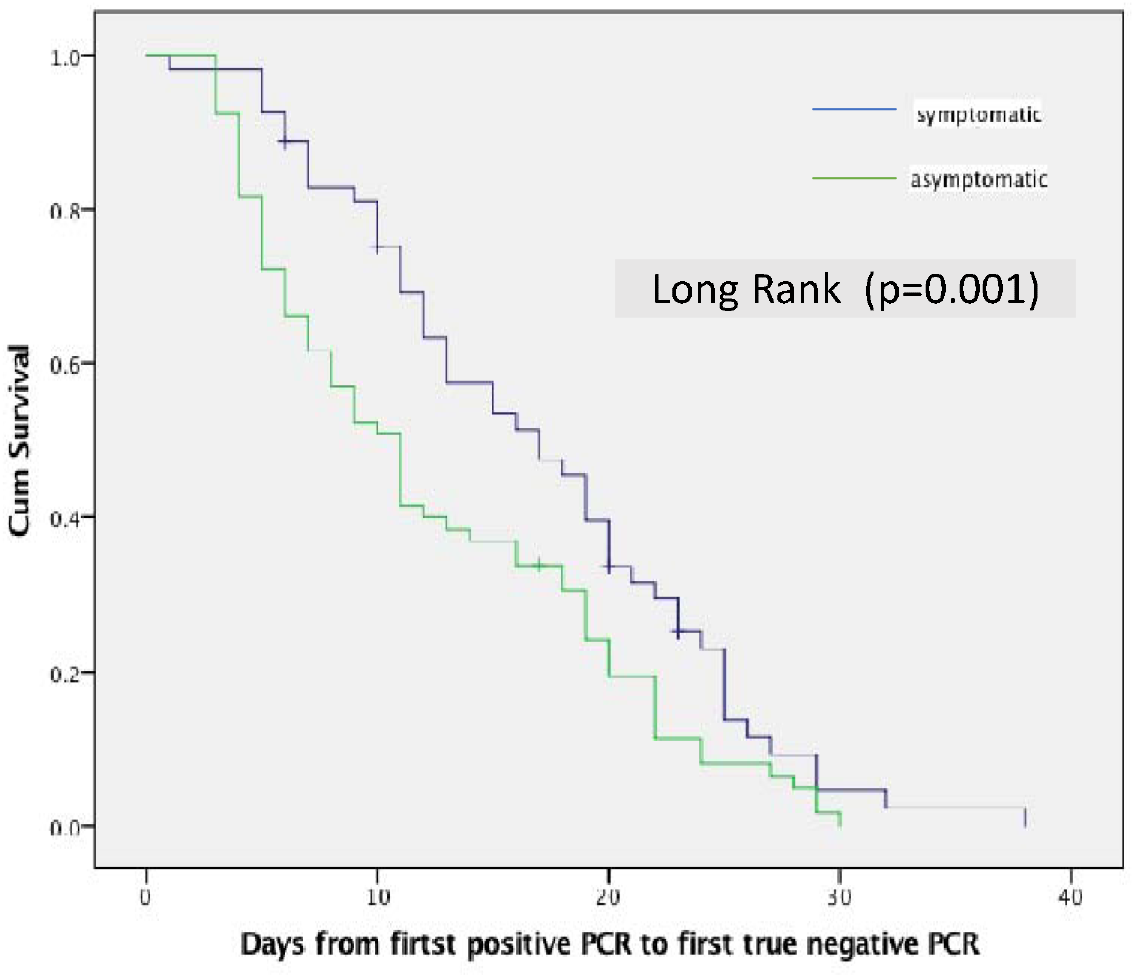
Kaplan Meier curve comparing the median time to virus clearance among the symptomatic and asymptomatic group

## Discussion

The first confirmed case of SARS-COV-2 in Saudi Arabia was on March 2^nd^, 2020, for a Saudi traveler who came back from Iran. In response to the COVID19 pandemic, Saudi Arabia was one of the first countries that applied early and very strict mitigation measures while the number of confirmed cases was still low (300 cases)^8^. On March 15^th^, 2020, the Saudi CDC (Weqaya) imposed a new regulation for all travelers coming from outside the kingdom, which is to be quarantined for 14 days in a designated facility regardless of the presence or absence of symptoms. This regulation has helped us to better understand the prevalence of SARS-COV-2 infection in asymptomatic patients.

In our cohort, the majority of residents were either truly asymptomatic (54%) or had mild symptoms (46%). The proportion of asymptomatic carriers in our cohort is considered to be one of the highest reported so far. Our proportion of asymptomatic carriers was somehow close to the reported proportion of the Diamond Princess cruise ship passengers in Japan (50%)^9^. However, a lower prevalence has been reported in other studies. A report from Germany on 116 travelers coming back from Wuhan city to Frankfort has shown a prevalence of 1.7% ^10^.

The most common symptoms were sudden and complete loss of taste and smell occurring in almost half of them (47.5%). More interestingly, these symptoms were found to be the only presenting symptoms in 18.7% of our residents, and this in itself should be considered as a red flag and should be reported as one of the common symptoms of mild or silent disease. Anecdotal reports of loss of taste and symptoms associated with COVID-19 have been reported. However, these symptoms have not been systematically addressed in studies. The majority of our symptomatic patients report that they could not smell even strong odors like perfumes and could not differentiate between sweet and sour food. Our findings are in light with a recent study that reported a 59% prevalence of loss of taste and smell in a cohort of COVID-19 patients^11^.

Most of the residents who were quarantined because of contact with lab-confirmed cases were identified by contact tracing programs. Although this information might be subjected to recall bias, none of our patients with a history of direct contact with lab-confirmed SARS-COV-2 infection (including household contacts) reported exposure to symptomatic individuals. Therefore, it indicates that they probably contracted the infection from asymptomatic or pre-symptomatic individuals. However, our study cannot confirm these findings.

Our study is unique in terms of the numbers of PCR performed per patient and also unique in terms of frequently testing the asymptomatic residents. A median of 5 tests per resident was done for both groups. This has allowed us to assess the virus clearance time accurately and to shed light on some of the PCR test short comes. Because we have enrolled asymptomatic residents in our study, we elected to use the first available positive PCR as the starting point for virus clearance time calculation. Keeping in mind that those asymptomatic residents might have contracted the virus a few days or weeks before the quarantine. Although our symptomatic group had mild symptoms, they have shed the virus for a longer period of time when compared with the asymptomatic group (17 vs. 11 days). Nevertheless, we had a few outliers. We had a 32 years old asymptomatic lady who cleared the virus after 36 days of admission. The clinical implications for persistently positive PCR in true asymptomatic and mild symptomatic patients need to be further studied. As a PCR test will not differentiate between a person who is contagious from a person who is not, further studies utilizing virus cultures and neutralization assays to help dictate the best duration of quarantine.

Our study result is different than what was reported by Lui et al. in which he found that majority of patients with mild symptoms cleared the virus at 10 days, while those with severe symptoms lasted for more than 10 days^12^. A false-negative test in a previously positive patient was not an uncommon finding in our cohort. This is most likely related to sampling techniques and specimen source. So, if the negative test was not repeated, we would have 18.8% of the time mistakenly reported these results as negative. So, we probably need to always do a repeat confirmatory test before deeming that the patient is infection-free. We also had about 3% of false-positive test results, which might be secondary to the detection of dead virus RNA particles rather than re-infection, as described recently by the South Korea group.

Although we have studied a unique population with a high prevalence of asymptomatic carriers, our study has few limitations. First, our study results cannot be generalized on all COVID-19 patients as we have excluded some high-risk groups. Second, our study is a cross-sectional study, so symptoms ascertainment before admission was subjected to recall bias. However, we find that majority of patients were well aware of their symptoms, and they were able to give us the date of onset of symptoms precisely.

**In conclusion**, the proportion of true asymptomatic carriers in our study was higher than reported. These findings re-enforce the importance of using test-based strategies and contact tracing, rather than symptoms-based screening checklist when dealing with travelers and subjects with direct contact to a lab-confirmed case. This testing-based screening strategy perhaps should be extended to first-line healthcare workers to limit the spread of the disease in hospitals. In addition, countries planning for mitigation measures release should enhance their testing abilities to prevent new outbreaks. Sudden onset of loss of smell and taste might be a predictor of having mild disease. These symptoms might as well be utilized as a marker for testing patients who may be positive for the virus.

## Data Availability

No supplementary materials are attached

## Notes

### Competing Interest Statement

The authors have declared no competing interest.

### Clinical Trial

This is neither a clinical trial nor a prospective study

### Funding Statement

This study is not funded

### Author Declarations

IRB of King Fahad Specialist Hospital- Dammam

### Summary of Updates

I made small changes in the title

